# Impact of self-administered pulse oximetry among non-hospitalized patients at risk of severe COVID-19 in Honduras: A pragmatic, block-randomized trial

**DOI:** 10.1101/2025.04.23.25326278

**Authors:** Kathryn W. Roberts, Berta Alvarez, Michael de St. Aubin, Omar Diaz, Salomé Garnier, C. Daniel Schnorr, Saul Cruz, Lorenzo Pavon, Angela Ochoa, Shiony Medice, Homer Mejía Santos, Yisela Martinez, Jonatán Ochoa, Sogeiry Solis, Devan Dumas, Margaret Baldwin, Alcides Martinez, Eric Nilles

## Abstract

The World Health Organization recommends remote monitoring and self-administered pulse oximetry to identify silent hypoxia and the need for medical intervention in non-hospitalized high-risk COVID-19 patients. These interventions have been evaluated previously, but evidence is needed to determine the impact on morbidity and mortality, particularly in lower- and middle-income countries.

**Methods:** A prospective, pragmatic, open-label trial was conducted in Tegucigalpa and Comayagüela, Honduras to evaluate the impact of self-administered pulse oximetry to reduce morbidity and mortality among non-hospitalized patients at high risk of adverse COVID-19 outcomes enrolled in a remote monitoring program. Participants were block-randomized to remote monitoring plus self-administered pulse oximetry versus remote monitoring alone. Participants in the pulse oximetry arm received daily calls to assess for high-risk clinical features, including hypoxia (oxygen saturation: SpO_2_ ≤94%) All participants reporting high risk symptoms were referred for in-person evaluation. The clinical trial is registered at https://clinicaltrials.gov/ct2/show/NCT04886414.

**Findings:** Between March 30, 2022 and January 24, 2023, 1,821 participants met the intention to treat analysis criteria, of whom 925 were randomized to remote monitoring and 897 to remote monitoring with pulse oximetry. Nearly ninety-nine percent of participants reported receiving one or more COVID-19 vaccine doses, and 90.2% three or more doses, with similar coverage across arms. Participants in the pulse oximetry arm were more likely to be referred for clinical evaluation (OR 1.60 [95% CI 1.09 – 2.46], p = 0.018), but not more likely to be hospitalized (OR 1.55 [95% CI 0.55 – 4.37, p = 0.401]. One participant died, two required intensive care, and none required mechanical ventilation; given limited data, these outcomes were not assessed.

**Interpretation:** Findings suggest that the use of self-administered pulse oximetry increased referral for additional care but did not result in different rates of hospitalization among a high-risk, but highly vaccinated, population with low rates of severe COVID-19. Given the infrequent progression to severe COVID-19, this trial did not assess whether self-administered pulse oximetry is related to mortality, need for mechanical ventilation, or admission to intensive care.

**Funding:** The U.S. Centers for Disease Control and Prevention funded the trial and supported design, interpretation, and review. All decisions were taken by the primary investigator. Rapid antigen tests were donated by Roche Diagnostics, Ltd, which had no additional involvement.

## Introduction

During the COVID-19 pandemic, healthcare facilities in Honduras were overwhelmed, and the Secretariat of Health (SESAL) needed strategies to provide quality care to high-risk, non-hospitalized patients. In 2020, the World Health Organization (WHO) developed guidelines to inform COVID-19 patient discharge and monitoring.(1) The guidance focused on health status, access to supportive care and monitoring, and prevention of onward transmission to caregivers. Building on this guidance, SESAL developed the *Practical Guide for the Home-based Management of Suspected or Confirmed COVID-19 Patients* to address the needs of non-critically ill patients at high risk for adverse outcomes in Honduras.(2) These guidelines recommend remote monitoring during the acute phase of SARS-CoV-2 infection to identify the need for medical intervention; evidence to inform the inclusion of pulse oximetry was needed.

Remote patient monitoring and self-administered pulse oximetry have been evaluated for COVID-19 and other diseases, but rarely in lower-middle income countries (LMIC).(3–7) Reviews of remote monitoring in LMIC during COVID-19 found it can improve healthcare accessibility, reduce health facility and provider burden, the risk of healthcare worker infections, and costs.(3,8) Concerns expressed about remote monitoring in LMIC highlight cost; lack of access to technology, mobile and electricity networks; and low technology literacy, especially among members of marginalized groups.(8–14) Additionally, pulse oximeters have been found to measure SpO_2_ 1.1 – 1.2% lower among people with darker skin, like people of Latine and Black descent in Honduras, which could delay referral to appropriate care.(15)

Silent hypoxia, low blood oxygen levels without dyspnea, can occur in COVID-19 patients prior to decompensation and may be associated with worse clinical outcomes; regular pulse oximetry may identify hypoxia before symptoms arise.(16–18) The WHO recommends self-administered pulse oximeter use “in symptomatic patients with COVID-19 and risk factors for progression to severe disease who are not hospitalized”.(19) If pulse oximeters are available, accurate, and used correctly, self-administered pulse oximetry may be a valuable addition to remote monitoring, according to a 2022 systematic review, although only 1 of 13 studies included was conducted in an LMIC (Brazil).(18) A systematic review of the safety and effectiveness of pulse oximeters for remote COVID-19 monitoring found that use is safe, reduces healthcare system burden, and identifies the need for care escalation, but did not demonstrate improved patient outcomes.(18) Recent retrospective studies have shown conflicting results, with one in the UK reporting a 52% lower odds of death among those with pulse oximeters, while a review in South Africa did not identify an association between self-administered pulse oximetry and reduced mortality.(20,21) A randomized trial in the United States that employed text message and phone-based remote monitoring did not demonstrate a difference in survival among patients randomized to receive a pulse oximeter versus those who were not.(22)

As of August 16, 2023, Honduras reported 474,566 cases of COVID-19 to the WHO, including 11,127 deaths (2.3% case fatality rate), with 112.18 deaths per 100,000.(23) COVID-19 vaccinations were introduced in March 2021, and as of March 2023, 65.85% of the national population was reported to have received at least one vaccine dose.(24) In an assessment of existing evidence, the U.S. Centers for Disease Control and Prevention (CDC) found that the likelihood of adverse COVID-19 outcomes, like hospitalization and death, was greater among older adults, people with certain underlying medical conditions, and people who experience barriers to healthcare access.(25) This study relied on that assessment to determine which patients were at high risk for adverse outcomes, and therefor eligible for enrollment.

This is the first randomized trial to examine the effectiveness of self-administered pulse oximetry to reduce mortality of COVID-19 patients at high risk for adverse outcomes in an LMIC. Other large-scale studies have examined patient outcomes retrospectively, but participants were not randomized to an intervention. Where participants were randomized to an intervention, the scale was insufficient to draw generalizable conclusions, and the studies were not conducted in LMIC.

The aim of this trial was to investigate the utility of including self-administered pulse oximetry in remote monitoring to reduce morbidity and mortality among non-hospitalized people with COVID-19 at high risk of adverse outcomes.

## Methods

### Study design and context

A prospective, pragmatic, block-randomized, open-label trial evaluated the effectiveness of remote monitoring versus remote monitoring with self-administered pulse oximetry between March 30, 2022 and January 24, 2023. After providing written informed consent, participants were enrolled by study-employed physicians between 7 am and 7 pm seven days per week at four SESAL-operated COVID-19 triage centers and one hospital-based COVID-19 screening site operated by the Honduran Social Security Institute in Tegucigalpa and Comayagüela, Honduras.

### Participants

Patients were eligible to participate in the trial if they reported COVID-19 symptoms, registered a positive SARS-CoV-2 Rapid Antigen Test (Roche, Switzerland) at the time of screening, lived or worked in Tegucigalpa or Comayagüela, had access to a functional telephone, were discharged home from the COVID-19 screening center by their treating physician, and were 60 years of age or older or 45-59 years of age with one or more U.S. CDC-defined high risk conditions.(25) High-risk conditions included cancer, cerebrovascular disease (stroke, transient ischemic attack), chronic kidney disease, chronic liver disease, chronic lung diseases (excluding asthma), diabetes mellitus type I or II, heart disease (heart failure, cardiomyopathy, coronary artery disease), mental health illness (depression, bipolar, or schizophrenia) tuberculosis, current or recent pregnancy, current or former smoker, and obesity. Gender categories were selected based on input from the Honduran study team and included female, male, non-binary, other (specify), and prefer not to disclose.

### Randomization and masking

Participants were block randomized on alternating days to remote monitoring, versus remote monitoring plus self-administered pulse oximetry. Intervention assignment was not masked.

### Procedures

COVID-19 screening center physicians were educated about the study protocol and referred patients to study staff for assessment; those who met inclusion criteria were invited to participate. Study-employed physicians administered informed consent, conducted enrollment interviews, and provided education about reducing household transmission. Participants assigned to receive an FDA-approved pulse oximeter (MedLine Soft Touch Fingertip Pulse Oximeter (Northfield, IL) or CuraPlex Fingertip Pulse Oximeter (Dublin, OH)) were educated on its use and instructed to record the lowest steady SpO_2_ upon waking, in the evening, and during the daily monitoring call in an SpO_2_ diary, which included pictorial and descriptive instructions. Additionally, participants received personal protective equipment to prevent disease transmission: N95 respirators for study participants, surgical masks for caregivers, bar soap, and alcohol-based hand rub (ABHR) was distributed to 50% of participants in both the remote monitoring and pulse oximetry arms. Analysis revealed no clinical effect associated with ABHR distribution; these findings are reported separately by the U.S. CDC Global WASH team.

Beginning the day after enrollment, study-employed nurses called participants daily. They administered a standardized questionnaire asking about warning signs or symptoms (hereafter referred to as warning signs), including blue/grey lips or nails, chest pain, difficulty breathing, new confusion, rapid breathing, or SpO_2_ ≤94% during a monitoring call, among those in the pulse oximetry arm. If any warning signs were reported, participants were referred for clinical evaluation at the site of enrollment. Participants were instructed to seek medical evaluation if warning signs arose outside of daily monitoring calls, including if their SpO_2_ reading was ≤94%. If SpO_2_ ≤94% was reported during a monitoring call, the participant received a referral to seek clinical evaluation. Referred participants were called the following day to ascertain whether they sought care and the outcome. Participants were registered as unreachable for the specified monitoring day if they and their secondary contact failed to answer a call after three attempts but were called again the next day. Participants were recorded as withdrawing from the study if they asked not to receive more calls. Participants were considered lost to follow-up if they did not complete a disenrollment interview and did not withdraw from the study. Phone-based monitoring was continued until ten days post symptom onset unless the participant was (i) admitted to a hospital for > 24 hours, at which point daily calls were discontinued, or (ii) reported subjective or measured fever (≥38.0 C) during the prior 24 hours in which case daily monitoring continued until they were fever-free for 24 hours.

To assess the outcomes of participants with warning signs who were referred for additional clinical evaluation, study staff reviewed charts at COVID-19 screening centers and hospitals using standardized data collection forms on electronic tablets. Forms included date of admission and discharge, level of care, discharge disposition, vital signs, results of laboratory and radiographic studies, medication received, and medical interventions such as supplemental oxygen administration. If study staff could not locate patient records, they called the participants to confirm the date and location of their visit. Medical records were recorded as missing if they could not be located.

### Outcomes

The primary trial outcome was the difference in mortality between study arms between March 30, 2022, and January 24, 2023. Secondary outcomes were risk of referral, SpO_2_% upon presentation for additional care, hospitalization, intensive care unit (ICU) admission, need for mechanical ventilation, and duration in ICU, and ventilator requirement.

Adverse events and safety were assessed by the study team during daily monitoring phone calls with participants; any concerns were brought to the trial team for review.

### Statistical analysis

Sample size was calculated using a two-sided test of independent proportions with the statistical parameters of alpha 0.05 and 0.8 power. Estimates of anticipated adverse outcomes were based on data provided by SESAL in November 2021 from the most recent COVID-19 wave, attributed to the Delta variant. Sample size calculations were conducted individually for the primary outcome and secondary outcome. The largest target sample size was selected as the trial target (n=1,862).

The analysis employed an intention-to-treat approach; all participants who met inclusion criteria and were randomized to an intervention were included in the analysis. Two-sided t-tests were used to compare the proportions of participants experiencing primary and secondary outcomes between study arms. Median with interquartile range (IQR) or mean with standard deviation (SD) are used to present descriptive statistics. Univariable logistic regression was used to estimate the odds ratio (OR) of outcomes between those with versus without pulse oximetry. Multivariable regression models included all variables with p<0.10 significance during univariate analysis, to ensure inclusion of variables that show lower significance in univariate regression but may become significant in multivariable analysis. Variables were considered significant within the multivariable models if p<0.05. Differences were analyzed to determine odds ratios, and confidence intervals were calculated with 95% confidence. Data were analyzed using STATA (version 17.0) and R (version 4.2.2).(26,27)

A Data and Safety Monitoring Board was not established, given the intervention was considered low risk. The team implementing the trial continuously supervised safety and conducted several interim analyses. The clinical trial is registered at https://clinicaltrials.gov/ct2/show/NCT04886414.

## Results

Between March 31, 2022, and January 15, 2023, study staff screened 1,860 patients with a positive SARS-CoV-2 antigen test; 1,821 were enrolled (97.9%) and included in the intention-to-treat analyses. Of these, 924 (50.7%) were randomized to remote monitoring and 897 (49.3%) to remote monitoring plus pulse oximetry. Of the 1,821 participants, 1,768 (97.1%) completed study monitoring per protocol including 900/924 (97.4%) in the remote monitoring arm and 868/897 (96.8%) in the pulse oximetry arm. Treatment arm assignment was similar across enrollment sites (p = 0.47). The trial profile **(Figure 1)** identifies the number of patients referred for enrollment (n=1,860), those who declined (n=19), did not meet inclusion criteria (n=19), were enrolled twice (n=1), withdrew (n=5), were lost to follow-up (n=48), were referred for additional care (n=100), and completed the discharge questionnaire per protocol (n = 1,768). Only participants in the remote monitoring arm withdrew (n=5), but the association was not statistically significant (p=0.062). Being lost to follow-up was not associated with the study intervention (n=48, OR 1.59 [95% CI 0.89 – 2.86], p=0.120). Chart review was attempted for all fifty-two participants referred for follow-up and who reported attending or when attendance was unknown. Study staff called participants if the record of their return for care could not be found at the health facility. During these calls, twenty-four (46.1%) participants changed their responses, reporting that, in fact, they never sought additional care. Of the remaining 28 participants, 22 (79%) medical records were located and reviewed, five (18%) could not be located, and one participant reported seeking care at a health facility outside of the study area.

**Figure 1.**
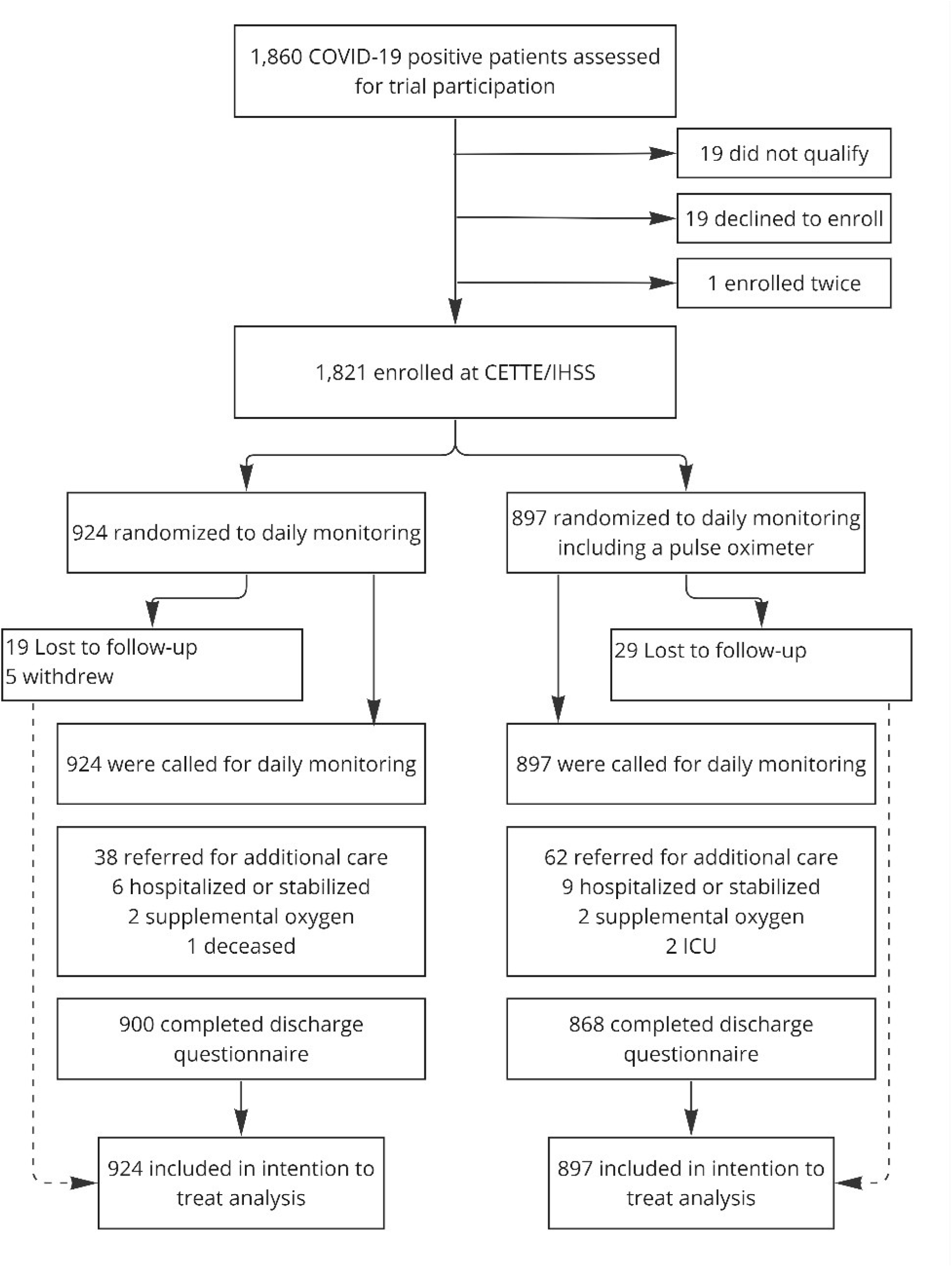
Trial profile.

The median participant age was 61 years (IQR 53-68), 1107/1,821 (60.8%) were female, and 1,665/1,821 (91.4%) reported at least one high-risk comorbidity. The most frequently reported comorbidities were hypertension (63.7%, n = 1,160), obesity (39.3%, n=716), and diabetes (30.2%, n=550). Sixty-one percent (61.6%, n= 1,123) of participants reported no previous SARS-CoV-2 infection. 98.9% (n=1,799) had received one or more COVID-19 vaccine doses, and 90.2% (n=1,160) had received three or more doses. Enrollment occurred with a mean of 2.9 days (SD 1.5) post-symptom onset. Participant characteristics were similar across treatment arms (**Table 1**).

**Table 1.** Baseline Characteristics Enrolled Patients In A Randomized Trial Of Self-Administered Pulse Oximetry Among Covid-19 Patients In Honduras.

| Characteristic | Treatment |  | Total |
| --- | --- | --- | --- |
|  | Without pulse oximeter | With pulse oximeter |  |
| **N** | 924 | 897 | 1,821 |
| **Age – Median (IQR) - years** | 61 (54-68) | 61 (53-69) | 61 (53-68) |
| **Gender n (%)** |  |  |  |
| **Female** | 565 (61.1%) | 543 (60.4%) | 1107 (60.8%) |
| **Male** | 358 (38.7%) | 353 (39.4%) | 711 (39.0%) |
| **Other** | 3 (0.1%) | 2 (0.0%) | 3 (0.015%) |
| **Ethnic group n (%)** |  |  |  |
| Participants selected all applicable groups |  |  |  |
| **Mestizo or Ladino** | 874 (94.5%) | 867 (96.7%) | 1741 (95.6%) |
| **Lenca** | 8 (0.9%) | 9 (1.0%) | 17 (0.9%) |
| **Other ethnicity** | 49 (5.1%) | 28 (3.0%) | 77 (4.2%) |
| **City of residence** |  |  |  |
| **Comayagüela** | 398 (43.1%) | 410 (45.7%) | 808 (44.4%) |
| **Tegucigalpa** | 526 (56.9%) | 487 (54.3%) | 1013 (55.6%) |
| **Education n (%)** |  |  |  |
| **No formal education** | 13 (1.4%) | 12 (1.3%) | 25 (1.4%) |
| **Any primary** | 221 (23.9%) | 246 (27.4%) | 467 (25.6%) |
| **Any secondary** | 337 (36.4%) | 305 (34.0%) | 642 (35.2%) |
| **Any university** | 350 (37.8%) | 330 (36.8%) | 680 (37.4%) |
| **Occupation n (%)** |  |  |  |
| **Homemaker** | 216 (23.4%) | 238 (26.5%) | 454 (24.9%) |
| **Active worker** | 518 (56.1%) | 476 (53.1%) | 994 (54.6%) |
| **Retired** | 162 (17.5%) | 155 (17.3%) | 317 (17.4%) |
| **Unemployed** | 28 (3.0%) | 28 (3.1%) | 56 (3.1%) |
| **Comorbidities n (%)** |  |  |  |
| **Chronic lung disease (excluding asthma)** | 27 (2.9%) | 25 (2.8%) | 52 (2.9%) |
| **Chronic kidney disease** | 13 (1.4%) | 10 (1.1%) | 23 (1.3%) |
| **Heart disease** | 45 (4.9%) | 48 (5.4%) | 93 (5.1%) |
| **Hypertension** | 583 (63.0%) | 577 (64.3%) | 1,160 (63.7%) |
| **Chronic liver disease** | 23 (2.5%) | 18 (2.0%) | 41 (2.3%) |
| **Diabetes** | 287 (31.0%) | 263 (29.3%) | 550 (30.2%) |
| **Tuberculosis** | 2 (0.2%) | 3 (0.3%) | 5 (0.3%) |
| **Cerebrovascular disease** | 24 (2.6%) | 26 (2.9%) | 50 (2.7%) |
| **Cancer** | 43 (4.6%) | 41 (4.6%) | 84 (4.6%) |
| **Psychiatric illness** | 85 (9.2%) | 72 (8.0%) | 157 (8.6%) |
| **Current or former smoker** | 217 (23.5%) | 208 (23.2%) | 425 (23.3%) |
| **Obesity (BMI ≥30)** | 363 (39.2%) | 353 (39.4%) | 716 (39.3%) |
| **Last SARS-CoV-2 infection n (%)** |  |  |  |
| **None** | 575 (62.2%) | 548 (61.1%) | 1,123 (61.6%) |
| **0 - 3 months ago** | 2 (0.2%) | 6 (0.7%) | 8 (0.4%) |
| **4 - 6 months ago** | 32 (3.5%) | 35 (3.9%) | 67 (3.7%) |
| **7 - 12 months ago** | 97 (10.5%) | 88 (9.8%) | 185 (10.2%) |
| **>12 months ago** | 210 (22.7%) | 206 (23%) | 416 (22.8%) |
| **Don't know/Prefer not to say** | 9 (1.0%) | 14 (1.6%) | 23 (1.3%) |
| **Days since SARS-CoV-2 infection, mean (SD)** | 477.7 (238.8) | 463.8 (347.9) | 470.8 (243.3) |
| **Vaccination status n (%)** |  |  |  |
| **0 doses** | 10 (1.1%) | 10 (1.1%) | 20 (1.1%) |
| **1-2 doses** | 79 (8.6%) | 80 (8.9%) | 159 (8.7%) |
| **3-4 doses** | 864 (90.4%) | 806 (91.0%) | 1341 (90.2%) |
| **Days symptom onset to enrollment, mean (SD)** | 3.0 (1.5) | 2.9 (1.4) | 2.9 (1.5) |
| **Vital signs at enrollment** |  |  |  |
| **Elevated blood pressure (>130/80)** | 438 (47.4%) | 418 (46.6%) | 856 (47.0%) |
| **Fast heart rate, >100 beats/ minute** | 106 (13%) | 100 (13%) | 206 (13%) |
| **Respiratory rate > 18 per min** | 352 (38.1%) | 310 (34.6%) | 662 (36.4%) |
| **SpO2 ≤ 94%** | 31 (3.4%) | 38 (4.2%) | 69 (3.8%) |
| **Temperature ≥ 38.0 C** | 12 (1.3%) | 12 (1.3%) | 24 (1.3%) |

One person reported non-binary gender (pulse oximeter intervention), one person reported “other” gender (no pulse oximeter), and three people preferred not to disclose their gender (two no pulse oximeter, one pulse oximeter); these have been aggregated into the other category.
<sup>2</sup> Maya, Garifuna, Negro Ingles, Tolupanes, Misquitos, Tawakas, prefer not to disclose ethnicity, and other ethnicity have been aggregated into the other category. All analyses were conducted using disaggregated ethnicity.

Most participants were enrolled during sequential waves of transmission June - August 2022 and December 2022 - January 2023. (**Fig 2**).

**Figure 2.**
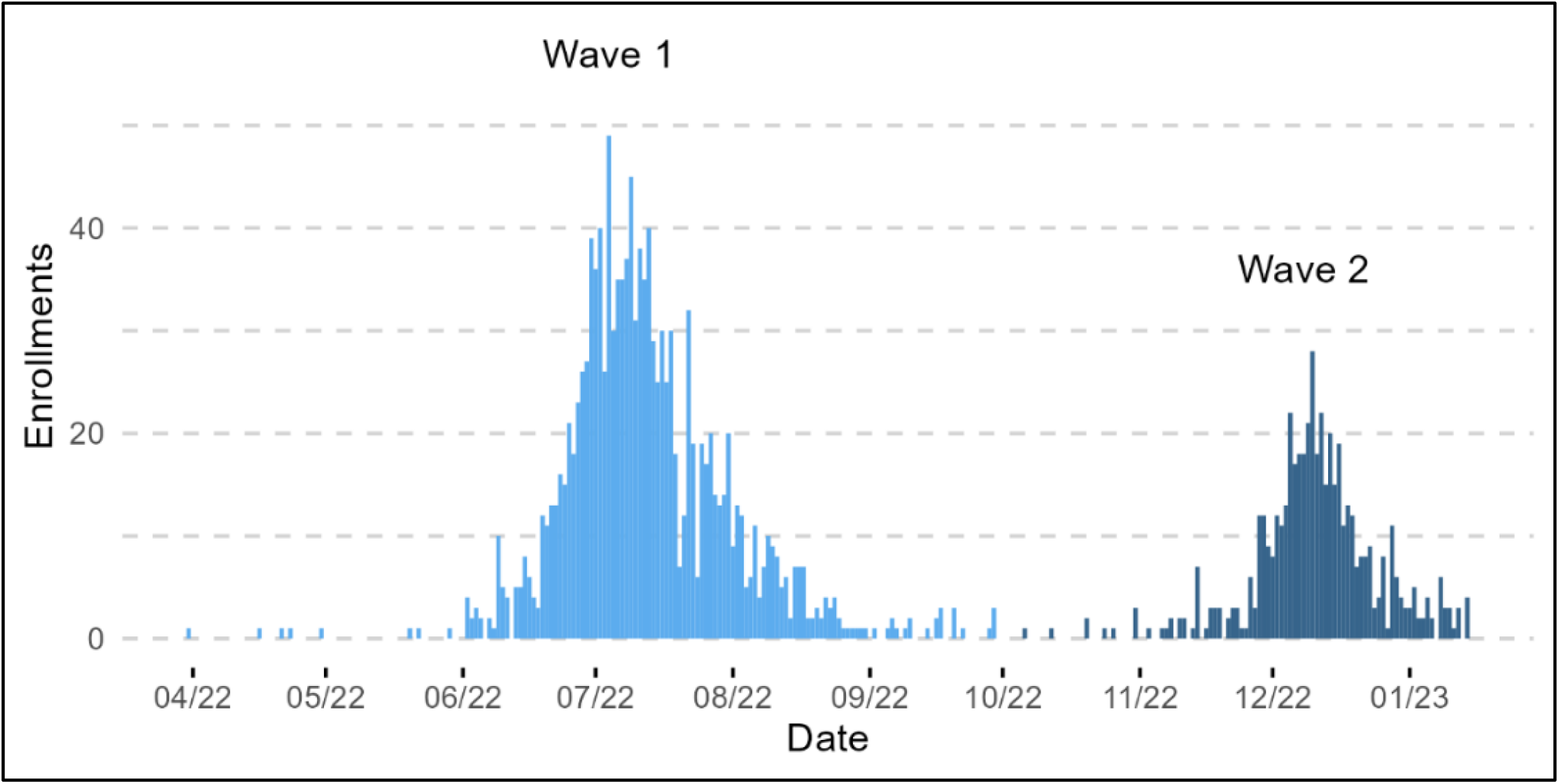
Daily Enrollment Of Covid-19 Patients In A Randomized Trial Of Self-Administered Pulse Oximetry In.

### Honduras

The primary outcome, the difference in mortality between the study arms, was not assessed due to low mortality (n=1).

A total of 12,307/12,602 (97.7%) daily monitoring calls were completed, with a mean of 6.9 (SD 3.0) completed calls per participant (**Table S1**). During monitoring calls, chest pain (2.4%, n=44) and difficulty breathing (1.3%, n=23) were the most commonly reported early warning signs. Over the course of the trial, 116 instances of self-reported warning signs were identified during 92 monitoring calls among 73 participants (4.0%). Among participants with a pulse oximeter, 315 (35.1%) reported at least one reading where SpO_2_ ≤94% at any of the three daily measurement timepoints; however, the reading was only stably ≤94% during a monitoring call among 47 (5.23%) participants, and therefore only resulted in a referral among those participants (**Table S1**). Enrollment in the pulse oximeter arm was associated with more frequent referrals, with 62 referrals (6.91%) versus 38 (4.23%) referrals among the remote monitoring arm (OR 1.73 [CI 1.14 – 2.62], p = 0.009). When excluding referrals due to low SpO_2_ (only measured in the intervention arm) there was no difference between the study arms (OR 0.82 [CI 0.51 – 1.31], p = 0.41), indicating pulse oximetry drove the increase in referrals in the intervention arm. Univariate analysis did not identify a statistically significant relationship between the likelihood of referral, COVID-19 vaccination status, COVID-19 wave during enrollment, age, or SpO_2_% at enrollment (**Table S2**). Variables identified as statistically significant in univariate analysis were incorporated into a multivariable model. Underweight body mass index (BMI <18.5) (OR 5.93, [95% CI 1.10 – 32.06]), and reporting chest pain (OR 3.04 [CI 1.76 – 5.23]), chills (OR 2.27, [CI 1.20 – 4.30]), diarrhea (OR 2.08 [CI 1.22 – 3.55]), or shortness of breath (OR 3.02 [CI 1.78 – 5.14]) at enrollment significantly increased the risk of later referral.

As part of the risk of referral analysis, the risk of ever recording SpO_2_ ≤94% at any of the three daily timepoints was analyzed among those in the pulse oximeter arm (n=315/897, 35.1%). Increased age (OR 1.03 [95% CI 1.01 – 1.05]), male gender, (OR 1.8 [CI 1.29 – 2.96]), obesity (BMI ≥30) (OR 2.03 [CI 1.32 – 3.13]), reporting fatigue at enrollment (OR 1.54 [CI1.02 – 2.32]), and reporting rapid respiratory rate during a remote monitoring call (OR 4.09 [CI 1.19 – 14.10]) significantly increased the odds of ever reporting an SpO_2_ ≤ 94% during a monitoring call (**Table S3**). For each percentage point higher the participant’s SpO_2_ was at enrollment, their odds of ever reporting hypoxia during the monitoring period were reduced by 0.75 (OR 0.75 [CI 0.66– 0.84]).

There difference in hospitalization rates between intervention arms was not statistically significant, with nine hospitalizations in the pulse oximetry arm versus six hospitalizations in the remote monitoring arm (OR 1.6 [95% CI 0.55 – 4.37], p=0.40]). Conversely, in multivariable analysis, residing in Tegucigalpa (rather than Comayagüela) (OR 5.18 [CI 1.07 – 25.04]) and reporting rapid respiratory rate during remote monitoring (OR 32.03 [CI 4.42 – 231.99]) were positively associated with risk of hospitalization (**Table S4, S5**). COVID-19 wave during enrollment, gender, or COVID-19 vaccination status were not associated with risk of hospitalization. The secondary outcomes of admission to the intensive care unit (n=2) and the need for mechanical ventilation (n=0) were not assessed due to limited outcome data.

### Adverse events

No adverse events or harms elated to remote monitoring or pulse oximeter use were reported.

## Discussion

In this pragmatic, block-randomized trial, we present data on the impact of including pulse oximetry in remote monitoring to reduce mortality among COVID-19 patients at high risk of adverse outcomes in Tegucigalpa and Comayagüela, Honduras. The findings did not demonstrate a difference in mortality between arms. However, the evidence is insufficient to assess the effectiveness of remote monitoring with versus without self-administered pulse oximetry. This trial was grounded in the concept that SARS-CoV-2 infection can cause silent hypoxia, and therefore, the measurement of oxygen saturation among non-hospitalized COVID-19 patients may lead to the earlier detection of hypoxia, leading to earlier intervention and better outcomes. In this trial, 315 (35.1%) of participants who received a pulse oximeter registered at least one reading where Sp02 ≤ 94%. Of these, 128 (40.6%) had an elevated respiratory rate at enrollment or reported rapid breathing during remote monitoring. This means that although lower SpO_2_% was associated with rapid respiratory rate at enrollment (OR 1.74 [95% CI 1.07 – 2.82], p<0.024]) and rapid breathing during remote monitoring (OR 3.59 [CI1.5 – 8.55], p = 0.004), pulse oximeters enabled the identification of silent hypoxia in 187 participants (59.4% of those who ever experienced hypoxia). However, perhaps unexpectedly, we did not identify a clinical benefit of identifying silent hypoxia among our study participants, likely because the degree of hypoxia among these individuals was mild with nearly all SpO_2_ readings below the 94% threshold between 90-93%.

Analysis of secondary outcomes demonstrated 1.6 times greater odds of referral among participants in the pulse oximetry arm, due to the inclusion of low SpO_2_% as a criterion for referral. There was no association between the intervention arm and other secondary outcomes. Across the entire study population, underweight BMI, reporting chest pain, chills, diarrhea, or shortness of breath at enrollment, were associated with increased referral during the monitoring phase. Older age, male gender, SpO_2_% ≤ 94% at enrollment, and reporting rapid respiratory rate during a monitoring call increased the odds of ever reporting hypoxia during the monitoring period. Underweight BMI has previously been associated with COVID-19 severity, including progression to pneumonia and development of secondary infections.(28) For each percentage point higher SpO_2_ at enrollment, the odds of reporting hypoxia during the monitoring period were reduced by 0.75. Among those referred who presented for care, SpO_2_% did not show a significant difference between interventions. Odds of hospitalization were not associated with the study intervention, however, residing in Tegucigalpa (rather than Comayagüela) and reporting rapid respiratory rate during remote monitoring were associated with increased risk of hospitalization. To better understand the association between city of residence and likelihood of hospitalization, we examined which variables were associated with city of residence and found a significant relationship between residing in Comayagüela and lower educational attainment (OR 1.17 [CI 1.05 – 1.30] p=0.004) and lower SpO_2_% at enrollment, whereby for each one unit increase in SpO_2_% at enrollment the odds that the participant lived in Comayagüela decreased by 0.879 [CI [0.816 – 0.946], p=0.001). Findings related to lower SpO_2_% at enrollment, and lower likelihood of hospitalization, could be related to differences in baseline health status, healthcare access, or healthcare seeking behavior.

The effectiveness of pulse oximetry to reduce morbidity and mortality among people with SARS-CoV-2 infections could not be determined in this trial, due to the low incidence of adverse outcomes among the trial population or because the intervention is ineffective, i.e. the null hypothesis could not be rejected. Other trials of self-administered pulse oximetry among COVID-19 patients have shown mixed results, with both positive and null results reported.(20,21) Differences in context, patient population, and concerns related to pulse oximeter accuracy must be considered when interpreting these findings. From our perspective, the body of evidence pertaining to self-administered pulse oximetry to reduce morbidity and mortality among COVID-19 patients at high risk for adverse outcomes does not yet meet the standard required to recommend this intervention for a broader population in this context, which includes high rates of previous infection, vaccination, and lower risk of severe disease with currently circulating SARS-CoV-2 variants. However, continuing to explore approaches to stratify patient risk and provide additional care to those most likely to experience morbidity and mortality is an essential approach for COVID-19 and other epidemic prone diseases, and bears further investigation.

### Limitations

Our study has some limitations. First, the limited incidence of adverse outcomes among study participants constrained our ability to draw conclusions about the primary and most secondary outcomes. Between the design and implementation of the study, vaccines became broadly available and uptake was high among study participants, population SARS-CoV-2 exposure continued to increase, and the dominant SARS-CoV-2 variant shifted from Delta to Omicron, known to cause less severe disease.(29) Second, inclusion criteria were minorly shifted after the study began. On May 5, 2022, five weeks after enrollment commenced, and after five participants had been enrolled (0.27% of the study population), inclusion criteria were expanded to align with the CDC’s updated recommendations, adding hypertension as a high-risk condition. Concurrently, age-related inclusion criteria were adjusted to include patients 45-59 with at least one high-risk condition or 60+, to better reflect age-related risk of adverse outcomes. This means that patients who would have been eligible for participation later in the trial were not invited to participate at the beginning of the trial; we are unable to determine the number of potential participants missed. Third, while every effort was made to extract data from participant charts, including calling each participant and their emergency contact, these paper records could not be located 17.8% (n=5) of the time, limiting our ability to determine the final disposition of these participants. Finally, despite pulse oximeters presenting a more objective view of current health status than self-reported symptoms, pulse oximeters have been found to measure SpO_2_ 1.1 – 1.2% higher than actual in Black and Latino patients with darker skin.(15) This could mean that there were study participants who were never referred despite experiencing hypoxia, and therefore, pulse oximeter utility may be limited in the study population without adjustment to definitions of hypoxia or access to more accurate pulse oximeters.

Future research directions to determine the effectiveness of pulse oximeters in reducing morbidity and mortality among COVID-19 patients at high risk for adverse outcomes likely would need to take place retrospectively, at the population level, to identify significant adverse outcomes to draw conclusions. This might, unfortunately, eliminate the possibility of intervention randomization, which was one of the strengths of this study.

This trial does not present evidence that supports the implementation of pulse oximeters alongside remote monitoring to reduce morbidity and mortality among people at high risk of adverse COVID-19 outcomes. As such, policy and practice recommendations about the inclusion of pulse oximetry alongside remote monitoring cannot be made. This study joins an inconclusive body of evidence on the impact of self-administered pulse oximetry to reduce mortality in different settings and with varying research methodologies.

## Data Availability

Data belong to the Secretariat of Health of Honduras and therefore are not publically available. They may be requested from Eric Nilles, who will work with SESAL to provide data to those making reasonable requests.

## Acknowledgements

The authors would like to thank the residents of Tegucigalpa and Comayagüela, Honduras who consented and participated in the study. We thank the nurses and doctors who worked tirelessly to implement these interventions. We thank the city, regional, and national members of the Honduras Secretary of Health and the Hospital Institute for Social Security.

## Data access statement

The data analyzed to prepare this manuscript are housed at the Secretariat of Health of Honduras and the Harvard Humanitarian Initiative and are available upon reasonable request to the corresponding author.

## Funding source

Funded by the U.S. Centers for Disease Control and Prevention, Cooperative Agreement Number U01GH002238 via Centers for Disease Control and Prevention, Central America Country Office, Guatemala; Reducing the morbidity and mortality due to acute febrile illnesses in Central America and the Dominican Republic. The trial was sponsored by the US CDC, which consulted on study design, analysis, and interpretation. All decisions were taken by the primary investigator.

Rapid antigen tests were provided through an Investigator Initiated Study with Roche Diagnostics, Ltd, which played no role in study design, data collection, analysis, interpretation, writing of the report, or publication.

## Ethical approval

This activity was determined to meet the definition of research [45 CFR 46.102(l)] involving human subjects [45 CFR 46.102 (e)(1)] and Institutional Review Board (IRB) review and was approved by Massachusetts General Brigham Institutional Review Board (2021P001143), the Autonomous University of Honduras (00003070), and SESAL. Secondary data analysis was approved by the Johns Hopkins Bloomberg School of Public Health Institutional Review Board (Approval No. 24586). The protocol was reviewed by the U.S. Centers for Disease Control and Prevention and determined to be research but the CDC was not engaged.

## Centers for Disease Control and Prevention (CDC) Disclaimer

The findings and conclusions in this report are those of the author(s) and do not necessarily represent the official position of the U.S. Centers for Disease Control and Prevention/the Agency for Toxic Substances and Disease Registry.

## Supplementary Materials

**Table 2: S1:**
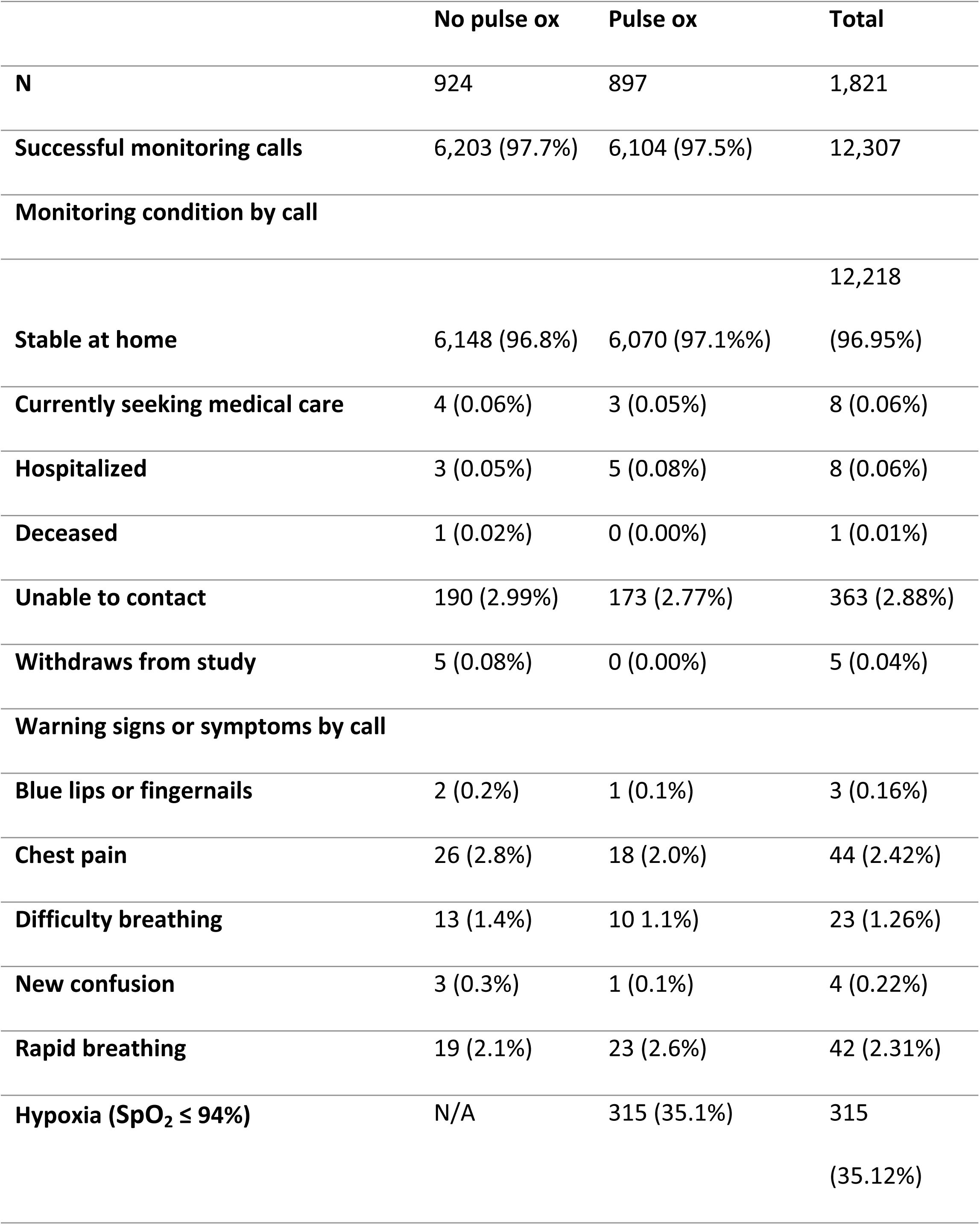
Monitoring calls and warning signs and symptoms.

**Table 3: S2:** Multivariable analysis of odds of referral for clinical assessment.

|  | OR (95% CI) | p value |
| --- | --- | --- |
| **Gender** |  |  |
| **Male** | *Ref* |  |
| **Female** | 1.32 (0.79 – 2.49) | 0.249 |
| **Health status** |  |  |
| **BMI – Healthy (18.6 – 24.9)** | *Ref* |  |
| **BMI – Underweight (<18.5)** | 5.93 (1.10 – 32.06) | **0.039** |
| **Psychiatric illness\*** | 1.48 (0.76 – 2.92) | 0.251 |
| **Symptoms at enrollment** |  |  |
| **Abdominal pain** | 1.00 (0.56 – 1.79) | 0.998 |
| **Chest pain** | 3.04 (1.76 – 5.23) | **0.000** |
| **Chills** | 2.27 (1.20 – 4.30) | **0.012** |
| **Diarrhea** | 2.08 (1.22 – 3.55) | **0.007** |
| **Dizziness** | 1.42 (0.68 – 2.93) | 0.348 |
| **Ear pain** | 1.15 (0.49 – 2.70) | 0.741 |
| **Fever (measured)** | 0.89 (0.46 – 1.71) | 0.719 |
| **Fever today (perception)** | 1.46 (0.80 – 2.69) | 0.218 |
| **Headache** | 1.80 (0.81 – 4.00) | 0.150 |
| **Muscle pain** | 0.84 (0.44 – 1.62) | 0.608 |
| **Nausea** | 1.57 (0.88 – 2.80) | 0.123 |
| **Shortness of breath** | 3.02 (1.78 – 5.14) | **0.000** |
| **Loss of smell** | 0.73 (0.33 – 1.61) | 0.437 |
| **Loss of taste** | 1.56 (0.76 – 3.21) | 0.227 |
| **Vomiting** | 1.42 (0.53 – 3.86) | 0.487 |

**Table 4: S3:** Multivariable analysis of odds of hypoxia during monitoring period.

|  | Odds ratio (95% CI) | p value |
| --- | --- | --- |
| **Demographics** |  |  |
| Age | 1.03 (1.01 – 1.05) | **0.000** |
| Gender, male | 1.75 (1.29 – 2.37) | **0.000** |
| **BMI** |  |  |
| Healthy weight (18.6 – 24.9) | Ref |  |
| Underweight (<18.6) | 1.60 (0.41 – 6.29) | 0.501 |
| Overweight (25 - 29.9) | 1.34 (0.89 – 2.02) | 0.164 |
| Obesity (≥30.0) | 2.03 (1.32 – 3.13) | **0.001** |
| **Comorbidities** |  |  |
| Heart disease | 1.3 (0.67 – 2.51) | 0.434 |
| **Previous COVID-19 infection** |  |  |
| No previous infection | Ref |  |
| ≤ 3 months | 0.80 (0.12 -5.43) | 0.818 |
| 4 – 6 months | 1.39 (0.66 – 2.91) | 0.386 |
| 7 – 12 months | 1.62 (1.00 – 2.64) | 0.051 |
| >13 months | 0.98 (0.68 – 1.41) | 0.911 |
| **Triage vital signs** |  |  |
| SpO<sub>2</sub> % | 0.75 (0.66 – 0.84) | **0.000** |
| Respiratory rate | 1.01 (0.92 – 1.11) | 0.832 |
| **Symptoms at enrollment** |  |  |
| Cough | 1.28 (0.77 – 2.15) | 0.345 |
| Fatigue | 1.54 (1.02 – 2.32) | **0.040** |
| Muscle pain | 0.73 (0.53 – 1.00) | **0.048** |
| **Monitoring warning signs** |  |  |
| **Chest pain** | 1.04 (.25 – 4.40) | 0.953 |
| **Rapid respiratory rate** | 4.09 (1.19 – 14.10) | **0.026** |
| **Difficulty breathing** | 3.42 (0.48 – 24.45) | 0.222 |

**Table 5: S4:**
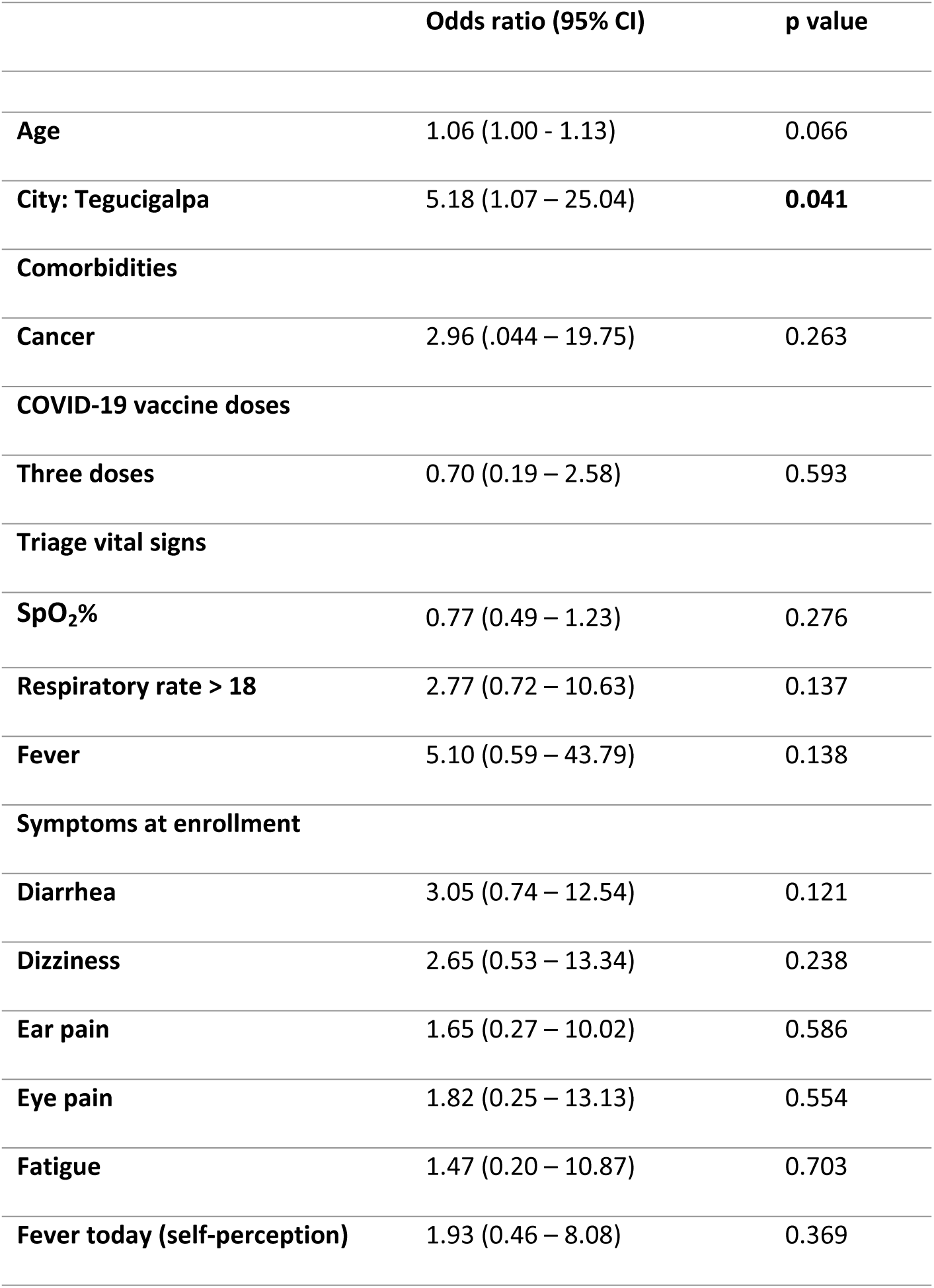
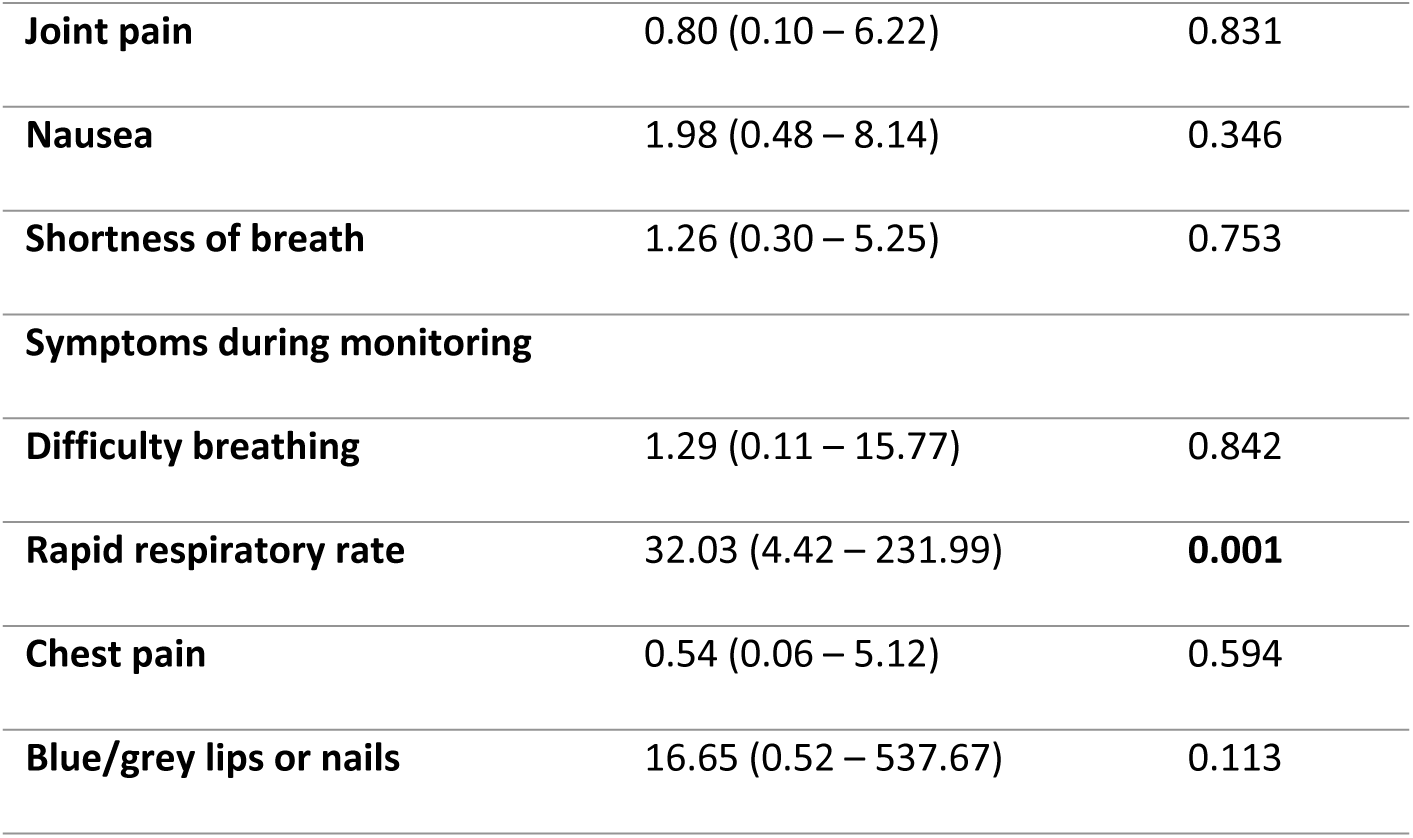
Multivariable analysis of odds of hospitalization.

**Table 6: S5:**
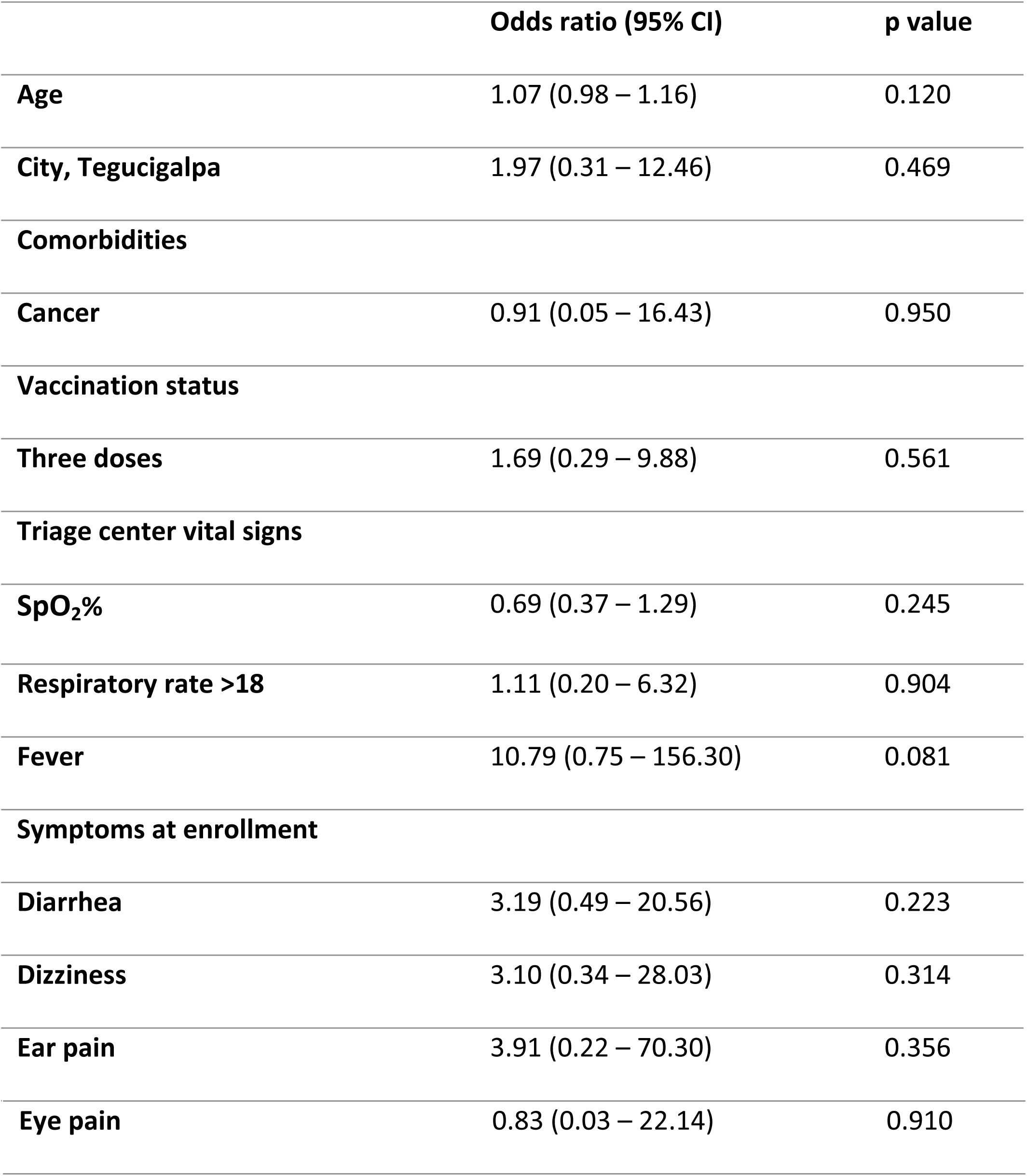
Multivariable analysis of odds of hospitalization, pulse oximeter arm only.

|  | **Odds ratio (95% CI)** | **p value** |
| --- | --- | --- |
| **Age** | 1.07 (0.98 – 1.16) | 0.120 |
| **City, Tegucigalpa** | 1.97 (0.31 – 12.46) | 0.469 |
| **Comorbidities** |  |  |
| **Cancer** | 0.91 (0.05 – 16.43) | 0.950 |
| **Vaccination status** |  |  |
| **Three doses** | 1.69 (0.29 – 9.88) | 0.561 |
| **Triage center vital signs** |  |  |
| **SpO<sub>2</sub>%** | 0.69 (0.37 – 1.29) | 0.245 |
| **Respiratory rate >18** | 1.11 (0.20 – 6.32) | 0.904 |
| **Fever** | 10.79 (0.75 – 156.30) | 0.081 |
| **Symptoms at enrollment** |  |  |
| **Diarrhea** | 3.19 (0.49 – 20.56) | 0.223 |
| **Dizziness** | 3.10 (0.34 – 28.03) | 0.314 |
| **Ear pain** | 3.91 (0.22 – 70.30) | 0.356 |
| **Eye pain** | 0.83 (0.03 – 22.14) | 0.910 |

